# Clinical and molecular analysis of a Chikungunya Virus outbreak in Southern Spain

**DOI:** 10.1101/2025.07.03.25330749

**Authors:** Maria Lara, Luis F. Caballero-Martínez, Irene Pedrosa-Corral, Carlos S. Casimiro-Soriguer, Cristina Gómez-Camarasa, Francisco J. Chamizo-López, Jose M. Navarro-Marí, Nicola Lorusso, Virtudes Gallardo, Inmaculada de Toro-Peinado, Antonio Sampedro-Martinez, Begoña Palop-Borrás, José María Reguera-Iglesias, Joaquin Dopazo, Sara Sanbonmatsu-Gámez, Javier Perez-Florido, Mercedes Pérez-Ruiz

## Abstract

Chikungunya Virus (CHIKV), primarily endemic to parts of Africa and Europe, has recently emerged as a public health concern in new geographic regions. This study reports the cases detected in Andalusia, southern Spain, during 2023. A multidisciplinary approach was employed, combining epidemiological investigations, virological diagnostic methods and viral genome sequencing and bioinformatic analysis to confirm the cases and identify transmission dynamics. During 2023, 32 probable or confirmed imported human cases of CHIKV infection were investigated in Andalusia. More than 90% of the cases were visiting friends and relatives and immigrants from Paraguay. Female gender, a subacute or chronic infection and arthritis and skin rash were the most frequent clinical-epidemiological characteristics. Serological and/or molecular testing confirmed CHIKV as the causative agent. Phylogenetic analysis of the viral genomes corresponding to four, out of the nine confirmed cases, suggested close relatedness to strains currently circulating in Brazil and Uruguay. This study highlights the geographic expansion of CHIKV and underscores the need for enhanced surveillance and vector control strategies in Spain. Public health interventions should focus on preventing further human especially in areas where imported cases are detected, based on a one health approach that includes vector control and mosquito populations monitoring.

## Introduction

Chikungunya virus (CHIKV) is an arthropod-borne RNA virus in the *Alphavirus* genus of the family *Togaviridae*, transmitted primarily by *Aedes* mosquitoes, notably *Aedes aegypti* and *Aedes albopictus* (1). Since its identification in Tanzania in 1952, CHIKV has caused numerous outbreaks globally, characterized by acute febrile illness and severe polyarthralgia (2). In recent decades, Europe has witnessed an emergence of CHIKV, mainly attributed to travelers coming from endemic areas, the increase in international travel and the expanding range of *Aedes albopictus* mosquitoes in mainland Europe, have generated local transmission of the virus related to viremic travel-related cases in Italy and France since 2007 (3, 4). Spain, with its conducive climate, a significant flow of travelers from endemic areas, and established populations of *Aedes albopictus*, is susceptible to CHIKV local transmission (5).

The clinical presentation of CHIKV infection includes sudden onset of fever, joint pain, headache, and rash. Although the disease is generally self-limiting, joint pain can persist for months, leading to significant morbidity (6). Currently, there is no specific antiviral treatment or licensed vaccine for CHIKV, emphasizing the importance of vector control and preventive measures.

Genomic surveillance has been crucial in monitoring the evolution and spread of new emerging viruses In Andalusia (Southern of Spain), where a coordinated genomic surveillance circuit was established to systematically sequence and analyze genomes of emerging viral threats across the region (7). This circuit has been essential in the rapid analysis of the current CHIKV outbreak. This study examines a recent CHIKV outbreak in Southern Spain, analyzing epidemiological patterns, phylogenetic relationships to other recent outbreaks, and public health responses. Understanding these factors is crucial for enhancing surveillance, improving patient management, and implementing effective vector control strategies to mitigate the impact of future outbreaks.

## Materials and Methods

### Chikungunya cases

A total of 32 probable or confirmed CHIKV cases were treated in different hospitals and Health Centers in Andalusia, Southern Spain, during 2023. Samples were submitted to the Andalusian Virus Reference Laboratory for microbiological analysis. For laboratory diagnosis, we performed molecular assays on whole blood, serum and/or urine samples from all patients with acute disease (i.e less than 14 days since symptoms onset), and serological tests were conducted on serum samples from all patients following the procedures of the Surveillance Program in Andalusia (8).

For clinical and epidemiological analysis, we included patients aged 18 or older referred from Primary Care or the Emergency Department to the Tropical Diseases Consultation at Hospital Regional Universitario de Málaga (HRUM, Málaga, Andalusia, Spain) (9). We collected demographic (age, sex and relevant comorbidities), epidemiological (country of origin, travel status including immigrant or visiting friends or relatives (VFR)) and clinical data from medical records.

### Samples, molecular diagnosis and culture isolation

The Andalusian Protocol for CHIKV surveillance and alert, indicated that clinical samples from suspected cases of human CHIKV disease have to be referred to the Andalusian Virus Reference Laboratory (AVRL) and, following the laboratory and epidemiological criteria, the cases were classified as confirmed or probable (8). The following diagnostic tests were performed in order to confirm CHIKV infection (9, 10): detection of virus-specific IgM and IgG antibodies in serum by ELISA testing (Euroimmun, Lübeck, Germany) and detection of specific viral nucleic acids in serum, whole blood and/or urine by VIASURE Zika, Dengue & Chikungunya Real Time PCR Detection Kit (Certest Biotech SL, Zaragoza, Spain) or RealStar® Chikungunya RT-PCR Kit 2.0 (Altona Diagnostics GmbH, Hamburg, Germany). Nucleic acid extraction from clinical samples was performed by using QIAsymphony DSP virus/pathogen mini kit (Qiagen, Hilden, Germany). For virus isolation, all of the procedures were performed within certified biosafety cabinets under biosafety level 3 (BSL3) containment. CHIKV RNA-positive samples were inoculated onto confluent monolayers of Vero cells. Passage to fresh Vero cell tubes was performed after 10 days of incubation or when cytopathic effect was observed (11). Viral growth was confirmed by qRT-PCR of the cell culture supernatant and viral culture was considered negative after 2 passages without evidence of CPE and negative qRT-PCR of the supernatant.

### Viral sequencing

Nucleic acids were extracted from 200⍰μL of each viral isolate using the QIAsymphony DSP virus/pathogen mini kit (Qiagen, Hilden, Germany), with a final elution volume of 60⍰μL. First-strand, cDNA synthesis was performed using 15 μL of purified RNA with ProtoScript II First strand synthesis kit (New England Biolabs, Ipswich, MA, USA) followed by the NEBNext Ultra Non-Directional Second Strand Synthesis module (New England Biolabs) as recommended by TWIST Bioscience (San Francisco, USA). The resulting double-stranded cDNA was purified using Agencourt AMPure XP beads (Beckman Coulter) with a 1.2X bead-to-sample ratio. Fifty nanograms of the purified double-stranded cDNA was used as input for adapter ligation, indexing and pre-capture amplification using the DNA Prep with Enrichment (Illumina Inc), following manufacturer’s instructions. Pre-capture amplification was performed using 12 cycles. Indexed libraries were pooled by mass, adding 250 ng of each library. Hybridization was performed overnight for 16 hours using Twist Comprehensive Viral Research Panel (Twist Bioscience). Hybridisation targets were then captured with Streptavidin Binding Beads. Post-capture amplification was performed on the enriched libraries (8 cycles). Final enriched libraries were quantified with Qubit 4 fluorometer (Invitrogen). Sequencing was carried out on a MiSeq system using a MiSeq Reagent Kit V2 (300-cycles).

### Viral genomic data processing

Sequencing data (150 bp x 2) were analyzed using custom in-house scripts and the nf-core/viralrecon pipeline software (v.2.6.0) (12). Briefly, after quality filtering of the reads, sequences for each sample were aligned to the CHIKV genome (KP164569.1) (13) using the *Bowtie 2* algorithm (v.2.4.4) (14), followed by identification and marking of duplicate reads using the *Picard* [14] tools (v.3.0.0). Genomic variants were identified with *iVar* (15) software (v.1.4), employing a minimum allele frequency threshold of 0.25 for variant calling and applying a filtering step to keep variants with a minimum allele frequency threshold of 0.75. Finally, using the set of high confidence variants and the reference genome, a consensus genome per sample was constructed with *bcftools* (v.1.16).

### Phylogenetic analysis

A phylogenetic analysis was performed using the Augur application (v.27.0.0) (16), whose functionality relies on the IQ-TREE (v.2.4.0) software (17). The MAFFT program (v.7.515) (18), which is one of the most sensitive multiple alignment methods (19), was utilized for the multiple alignment, using the strain NC_004162.2 (20) as reference. The phylogenetic tree is recovered by maximum likelihood, using a general time reversible model with unequal rates and unequal base frequencies (21). Branching date estimation was carried out with the least square dating (LSD2) method (22) using TreeTime v.0.11.4 (23). Branching point reliabilities were estimated by UFBoot, an ultrafast bootstrap approximation to assess branch supports (24).

The results can be viewed in the Nextstrain local server (25), which is now part of the Andalusian Genomic Epidemiology System (SIEGA) (26).

## Ethics approval

The study protocol was conducted in accordance with the Declaration of Helsinki and the ethical considerations of epidemiological research. This was a non-interventional study, with no further investigation into procedures described in this work. Ethical approval was obtained from the Comité de Ética de la Investigación-Málaga (CEIm) with the reference number SICEIA-2025-001086, on May 29th, 2025.

## Results

### Clinical-epidemiological data of Chikungunya cases

From January to July 2023, 18 patients with confirmed or probable CHIKV infection (54.5% of investigated cases) were treated at HRUM. All of the patients, except for one person who had traveled to Peru, were from Paraguay. Patients’ demographic, epidemiological and clinical characteristics are described in Table 1. All five acute-onset patients had a positive molecular diagnosis of CHIKV infection, the rest being diagnosed based on serologic tests (positive IgM and IgG).

**Table 1.** The demographic, epidemiological, and clinical characteristics of the patients.

| Characteristic | class | N(%) |
| --- | --- | --- |
| Gender | Female | 14 (77.8) |
|  | Male | 4 (22.2) |
| Age (years) |  | 42 (34-55)<sup>1</sup> |
| Infection at admission | Acute | 5 (27.8) |
|  | Subacute / Chronic | 13 (72.2) |
| Paraguay Department | Capital District | 7 (38.9) |
|  | Central | 3 (16.7) |
|  | Cordillera | 2 (11.1) |
|  | Alto Paraná | 2 (11.1) |
|  | Caaguazú | 1 (5.6) |
|  | Concepción | 1 (5.6) |
| Perú | NA | 1 (5.6) |
| NA | NA | 1 (5.6) |
| Epidemiology | VFR | 13 (72.2) |
|  | Immigrant | 4 (22.2) |
|  | Tourist<sup>2</sup> | 1 (5.6) |
| Comorbidities | Hypertension | 3 (16.7) |
|  | Diabetes | 1 (5.6) |
|  | Dyslipidemia | 1 (5.6) |
|  | Bone and joint disease | 2 (11.1) |
| Symptoms at admission | Fever | 17 (94.4) |
|  | Headache | 4 (22.2) |
|  | Arthritis | 16 (88.9) |
|  | Skin rash | 14 (77.8) |
|  | Diarrhea | 6 (33.3) |
|  | Conjunctivitis | 3 (16.7) |
| Follow up | Three months | 16 (88.9) |
|  | Six months | 6 (33.3) |
| Symptoms at three months | Arthralgia | 8/16 (50.0) |
|  | Arthritis | 3/16 (18.8) |
| Symptoms at six months | Arthralgia | 1/6 (16.7) |
|  | Arthritis | 0 (0%) |
| Treatment | NSAIDs<sup>3</sup> | 18 (100%) |
|  | Oral steroids | 10 (55.6%) |

<sup>1</sup> Median (interquartile range)
<sup>2</sup> Case from Perú
<sup>3</sup> NSAIDs, non-steroidal anti-inflammatory drugs.

### Microbiological diagnosis

During 2023, 43 clinical samples (34 serum, 8 urine, and 1 whole blood) were received at the AVRL, corresponding to 23 probable and 9 confirmed cases. Several patients provided more than one type of sample. IgM antibodies against CHIKV were detected in all 23 probable cases, with CHIKV-specific IgG also present in 20 of them.

Of the 9 confirmed cases, 2 were diagnosed by IgG seroconversion in paired serum samples collected two weeks apart. In the remaining 7 cases, confirmation was achieved by detection of CHIKV RNA through RT-PCR in at least one clinical specimen. RT-PCR yielded positive results in serum (n=7), urine (n=3), and whole blood (n=1). Virus isolation in cell culture was successful in 4 of the 7 RT-PCR-positive cases. The isolates were obtained from clinical samples provided by patients attending at HURM (n=2), Hospital Comarcal de La Axarquía, Málaga (n=1), and Hospital Universitario Reina Sofía, Córdoba (n=1). All four cell culture isolates underwent genomic sequencing for further characterization.

### Sequencing and data collection

The viral isolates were subjected to sequencing, and the resulting raw data were processed as described in the “Materials and Methods” section. A total of 4 high quality whole genome sequences were obtained. The CHIKV sequences are available in the European Nucleotide Archive (ENA) database under the project identifier PRJEB88493, and accessions: ERS24011617, ERS24011618, ERS24011619, ERS24011620.

A total of 810 CHIKV complete or near-complete genomes were found in the GenBank repository (listed in Supplementary Table 1). All these genomes were downloaded and aligned, together with the 4 CHIKV genome sequences obtained in this work, using the MAFFT program (see Materials and Methods)

### Phylogenetic analysis

Figure 1 shows a detail of the phylogenetic tree including the current samples sequenced in this study and some of the genetically closest CHIKV from previous outbreaks, whose sequences are available. Interestingly, the closest relative CHIKV sequences were reported in Brazil (2/2023), and belong to a clade that apparently was originally from this country, but in 2023, some viruses, descendant from this one, were detected in Paraguay as well. Currently three clades are cocirculating in Paraguay, one of them, related to the most prevalent clade circulating in Brazil, is also related to the samples sequenced in this study. A phylogenetic tree including the CHIKV sequences reported in this study and the 810 CHIKV sequences listed in Supplementary Table S1, representing the evolution of all the available sequences, was reconstructed as described in Materials and Methods and depicted in Supplementary Figure 1.

**Figure 1.**
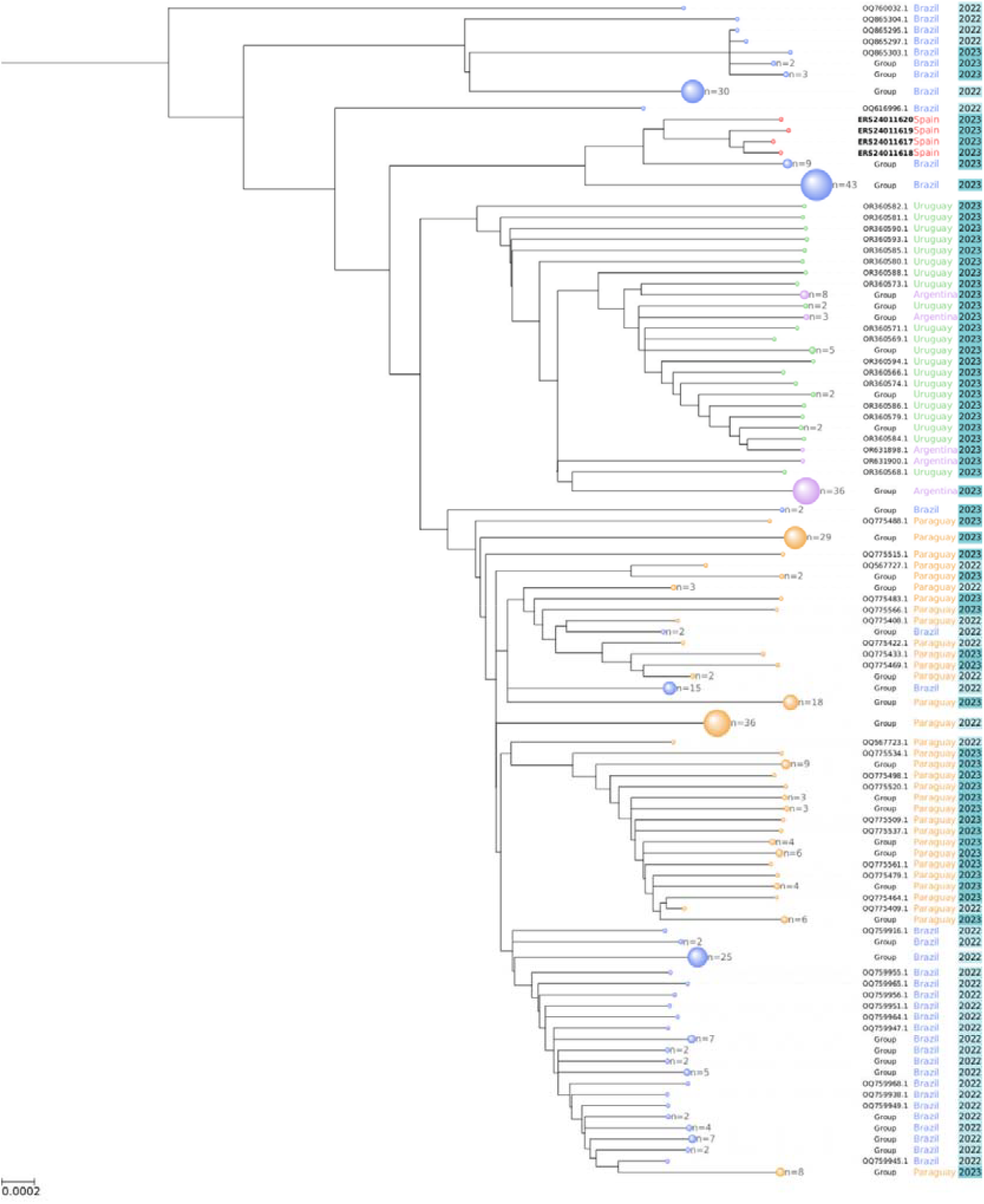
Detailed phylogenetic tree of the CHIKV sequences reported in this study in the context of the most closely related sequences available. The scale is nucleotide substitutions per site.

### The SIEGA Nextstrain server

The Andalusian Genomic Epidemiology System, SIEGA [25], includes a Nextstrain server [24] that offers an interactive view of the complete CHIKV phylogeny with the CHIKV viral genome sequences reported in this study in the context of some viral sequences of reference (see Supplementary Table 1). Supplementary Figure 1 shows a summarized view of the whole phylogenetic tree as displayed by the SIEGA Nextstrain server.

## Discussion

This work reports on a cluster of 32 cases of probable or confirmed CHIKV infection studied in Andalusia, Southern Spain, during 2023 (27). South America was the area of origin in 96.9% of cases, being 93.5% of them from Paraguay. This reflects the intense CHIKV transmission in South America, which experienced a major outbreak during 2022-2023 (28). Our findings highlight the importance of international travel in the importation of CHIKV cases into Europe and the potential for local transmission in receptive areas such as southern Spain, where *Aedes albopictus* is established (3).

Among the 18 cases with available clinical-epidemiological data, a clear predominance of cases in women was observed (77.8%), with a median age of 45 years (IQR 34-55), similar to previously reported series in the literature (29). Most patients were classified as VFR, reflecting the risk of imported diseases in this group, who normally relax the protective measures when going back to their country of origin and who represent a potential risk of local transmission in non-endemic countries when coming back from their travel. A majority of our cohort (72.2%) presented with subacute or chronic infection, aligning with previously reported trends in prolonged symptomatology following CHIKV infection (6, 30).

CHIKV infection also represents a substantial economic burden for non-endemic countries receiving travelers and VFR from endemic regions. The prolonged persistence of arthralgia and its impact on daily activities contribute to significant healthcare costs, work absenteeism, and social burden, including reduced productivity and the necessity for prolonged medical follow-up (29, 31). In our population, there was a significant proportion of patients who needed to continue follow-up at three months, with some of them even extending to six months. More than half of them had arthralgia lasting three months from the onset of symptoms and almost 20% of them had arthritis at that point, with these numbers decreasing when reaching the sixth month medical revision. This progressive decline in arthritis aligns with previously reported findings, which describe a similar resolution trend in CHIKV-associated arthritis over time (32).

We also support the role of nonsteroidal anti-inflammatory drugs (NSAIDs) as the cornerstone of arthralgia management, with oral corticosteroids being prescribed for more severe and persistent cases according to clinical practice guidelines (6). No other immunosuppressive therapy (i.e. methotrexate) was needed for the management of our patients.

From a public health perspective, this outbreak underscores the ongoing vulnerability of Europe to arboviral diseases. The risk of local transmission in Europe is raised by the presence of viremic individuals during these imported outbreaks, in regions where competent vectors with a broader ecological niche (i.e. *Aedes albopictus*) are established (2).

The genomic sequencing and phylogenetic analysis of four cases revealed that the viruses were closely related to strains circulating in Brazil and Uruguay, suggesting transcontinental transmission dynamics and emphasizing the need for continuous genomic surveillance. These findings are consistent with the known rapid dispersion of CHIKV facilitated by human mobility and the adaptability of the *Aedes* vectors (2).

As previously reported in a study using computational models, Mediterranean Europe is at a high risk of endemic transmission of CHIKV and other arboviruses (33), with our country, especially the southern and eastern regions such as Andalusia representing a vulnerable zone for local transmission of these infections. In this setting, the Andalusian surveillance system (SIEGA) and integrated genomic monitoring provide valuable tools to detect and trace such events promptly (26). Strengthening entomological surveillance, improving traveler screening, and educating healthcare professionals about early recognition and reporting are vital components of response strategies.

Similarly, genomic surveillance has demonstrated to be a key resource for accurately monitoring the evolution and spread of emerging viruses (34). Andalusia has implemented a genomic surveillance circuit (7) that was decisive in recent emerging viral threats, such as the SARS-CoV-2 pandemics (35, 36), the 2022 monkeypox outbreak (37), the successive occurrences of endemic and new West Nile Virus lineages (38, 39) and has been essential in the rapid analysis of the current CHIKV outbreak.

The strength of our study resides in the combination of clinical, epidemiological, and genomic data to understand viral introductions in our region. To the best of our knowledge, this is the only report of an imported outbreak of CHIKV infection in Europe related to the major outbreak in South America during 2022-2023. We also performed genomic sequencing and phylogenetic analysis which links our cases to those impacting the Americas in the outbreak, specifically the lineages reported in Brazil and Uruguay.

However, the study had some limitations, such as its retrospective nature, which limited the collection of patient data. We also limited the scope of our study to cases seen in our institution, which reduced the sample size and underestimated patients with milder disease who did not seek medical attention or were seen in primary care, thus underestimating the incidence of this imported outbreak in our country. In our study, we did not report the impact of musculoskeletal symptoms on patients’ quality of life, limiting our ability to adequately address this important issue.

In conclusion, this imported outbreak of CHIKV infection highlights the main clinical characteristics of this disease, especially the musculoskeletal symptoms during the follow up period. The detection of CHIKV among travelers, together with phylogenetic evidence linking these cases to recent South American outbreaks, reinforces the need for sustained vigilance in Europe due to new transmission hotspots and increased global mobility vigilance (40).

## Data Availability

The CHIKV sequences are available in the European Nucleotide Archive (ENA) database under the project identifier PRJEB88493, and accessions: ERS24011617, ERS24011618, ERS24011619, ERS24011620.

## Funding

This research was funded by grants PT17/0009/0006 and PMP24/00024 from the ISCIII, co-funded by the European Regional Development Fund (ERDF) as well as H2020 “ELIXIR-EXCELERATE fast-track ELIXIR implementation and drive early user exploitation across the life sciences” (GA 676559).

## Notes

### Competing Interest Statement

The authors have declared no competing interest.

### Author Declarations

The study protocol was conducted in accordance with the Declaration of Helsinki and the ethical considerations of epidemiological research. This was a non-interventional study, with no further investigation into procedures described in this work. Ethical approval was obtained from the Comite de Etica de la Investigacion-Malaga (CEIm) with the reference number SICEIA-2025-001086, on May 29th, 2025.

