## Supplementary figures and images for "Clinical and molecular analysis of a Chikungunya Virus outbreak in Southern Spain"

### Supplementary Figure 1

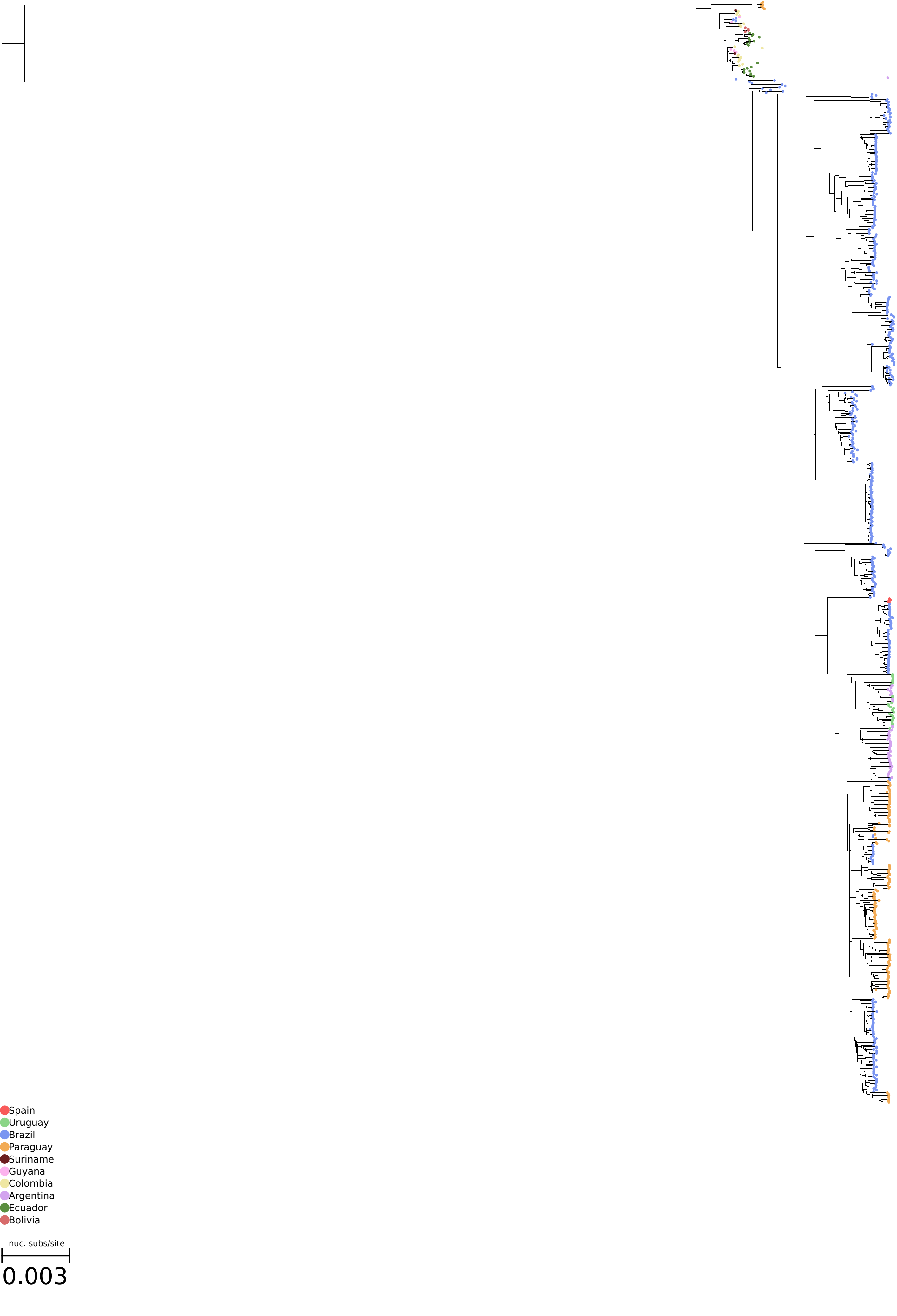
