## Supplementary Table 1 for "Clinical and molecular analysis of a Chikungunya Virus outbreak in Southern Spain"

**Supplementary Table 1**. complete or near-complete genomes were found in the GenBank repository

| Accession | Isolation date | Country | DOI | Pubmed ID |
|---|---|---|---|---|
| OR631892.1 | 02-2023 | Argentina | | |
| OR631865.1 | 02-2023 | Argentina | | |
| OR631867.1 | 02-2023 | Argentina | | |
| OR631916.1 | 05-2023 | Argentina | | |
| OR631899.1 | 04-2023 | Argentina | | |
| OR631917.1 | 04-2023 | Argentina | | |
| OR631898.1 | 04-2023 | Argentina | | |
| OR631897.1 | 04-2023 | Argentina | | |
| OR631900.1 | 04-2023 | Argentina | | |
| OR631895.1 | 04-2023 | Argentina | | |
| OR631894.1 | 03-2023 | Argentina | | |
| OR631891.1 | 03-2023 | Argentina | | |
| OR631896.1 | 03-2023 | Argentina | | |
| OR631901.1 | 03-2023 | Argentina | | |
| OR631910.1 | 03-2023 | Argentina | | |
| OR631908.1 | 03-2023 | Argentina | | |
| OR631907.1 | 03-2023 | Argentina | | |
| OR631906.1 | 03-2023 | Argentina | | |
| OR631909.1 | 03-2023 | Argentina | | |
| OR631889.1 | 03-2023 | Argentina | | |
| OR631884.1 | 03-2023 | Argentina | | |
| OR631864.1 | 03-2023 | Argentina | | |
| OR631879.1 | 03-2023 | Argentina | | |
| OR631882.1 | 03-2023 | Argentina | | |
| OR631886.1 | 03-2023 | Argentina | | |
| OR631866.1 | 03-2023 | Argentina | | |
| OR631883.1 | 03-2023 | Argentina | | |
| OR631881.1 | 02-2023 | Argentina | | |
| OR631887.1 | 02-2023 | Argentina | | |
| OR631863.1 | 02-2023 | Argentina | | |
| OR631893.1 | 02-2023 | Argentina | | |
| OR631890.1 | 02-2023 | Argentina | | |
| OR631885.1 | 02-2023 | Argentina | | |
| OR631868.1 | 02-2023 | Argentina | | |
| OR631880.1 | 02-2023 | Argentina | | |
| OR631888.1 | 02-2023 | Argentina | | |
| OR631869.1 | 02-2023 | Argentina | | |
| OR631872.1 | 02-2023 | Argentina | | |
| OR631870.1 | 02-2023 | Argentina | | |
| OR631871.1 | 02-2023 | Argentina | | |
| OR631873.1 | 02-2023 | Argentina | | |

| | | | | |
|---|---|---|---|---|
| OR631874.1 | 01-2023 | Argentina | | |
| OR631875.1 | 01-2023 | Argentina | | |
| OR631911.1 | 01-2023 | Argentina | | |
| OR631914.1 | 01-2023 | Argentina | | |
| OR631876.1 | 01-2023 | Argentina | | |
| OR631878.1 | 01-2023 | Argentina | | |
| OR631913.1 | 01-2023 | Argentina | | |
| OR631877.1 | 01-2023 | Argentina | | |
| OR631915.1 | 01-2023 | Argentina | | |
| MT150094.1 | 03-2015 | Bolivia | | |
| MT150098.1 | 05-2015 | Bolivia | | |
| MT150096.1 | 05-2015 | Bolivia | | |
| MT150093.1 | 03-2015 | Bolivia | | |
| KP164572.1 | 08-2014 | Brazil | 10.1186/s12916-015-0348-x | 25976325 |
| KP164567.1 | 08-2014 | Brazil | 10.1186/s12916-015-0348-x | 25976325 |
| KP164571.1 | 07-2014 | Brazil | 10.1186/s12916-015-0348-x | 25976325 |
| MN783353.1 | 10-2016 | Brazil | | |
| KU940225.1 | 07-2015 | Brazil | 10.1055/s-0036-1587323 | 27555980 |
| MT526900.1 | 01-2017 | Brazil | 10.3390/v12080853 | 32759878 |
| MG649980.1 | 04-2016 | Brazil | | |
| MG649979.1 | 03-2017 | Brazil | | |
| MG649981.1 | 07-2016 | Brazil | | |
| OL898663.1 | 09-2020 | Brazil | | |
| OL898714.1 | 11-2020 | Brazil | | |
| OL898669.1 | 12-2020 | Brazil | | |
| OL898675.1 | 12-2020 | Brazil | | |
| OL898668.1 | 01-2021 | Brazil | | |
| OL898707.1 | 01-2021 | Brazil | | |
| OL898682.1 | 01-2021 | Brazil | | |
| OL898689.1 | 01-2021 | Brazil | | |
| OL898690.1 | 01-2021 | Brazil | | |
| OL898674.1 | 01-2021 | Brazil | | |
| OL898700.1 | 01-2021 | Brazil | | |
| OL898696.1 | 01-2021 | Brazil | | |
| OL898704.1 | 01-2021 | Brazil | | |
| OL898664.1 | 01-2021 | Brazil | | |
| OL898683.1 | 01-2021 | Brazil | | |
| OL898667.1 | 02-2021 | Brazil | | |
| OL898676.1 | 02-2021 | Brazil | | |
| OL898710.1 | 02-2021 | Brazil | | |
| OL898670.1 | 02-2021 | Brazil | | |
| OL898711.1 | 02-2021 | Brazil | | |
| OL898699.1 | 02-2021 | Brazil | | |
| OL898706.1 | 02-2021 | Brazil | | |
| OL898697.1 | 02-2021 | Brazil | | |

| | | | | |
|---|---|---|---|---|
| OL898693.1 | 02-2021 | Brazil | | |
| OL898705.1 | 02-2021 | Brazil | | |
| OL898677.1 | 02-2021 | Brazil | | |
| OL898684.1 | 02-2021 | Brazil | | |
| OL898671.1 | 02-2021 | Brazil | | |
| OL898712.1 | 02-2021 | Brazil | | |
| OL898681.1 | 02-2021 | Brazil | | |
| OL898691.1 | 02-2021 | Brazil | | |
| OL898685.1 | 02-2021 | Brazil | | |
| OL898665.1 | 02-2021 | Brazil | | |
| OL898666.1 | 02-2021 | Brazil | | |
| OL898698.1 | 02-2021 | Brazil | | |
| OL898672.1 | 02-2021 | Brazil | | |
| OL898678.1 | 03-2021 | Brazil | | |
| OL898692.1 | 03-2021 | Brazil | | |
| OL898673.1 | 03-2021 | Brazil | | |
| OL898680.1 | 03-2021 | Brazil | | |
| OL898688.1 | 03-2021 | Brazil | | |
| OL898686.1 | 03-2021 | Brazil | | |
| OL898695.1 | 04-2021 | Brazil | | |
| OL898703.1 | 04-2021 | Brazil | | |
| OL898679.1 | 04-2021 | Brazil | | |
| OL898687.1 | 04-2021 | Brazil | | |
| OL898702.1 | 04-2021 | Brazil | | |
| OL898713.1 | 04-2021 | Brazil | | |
| OL898708.1 | 05-2021 | Brazil | | |
| OL898701.1 | 05-2021 | Brazil | | |
| OL898715.1 | 05-2021 | Brazil | | |
| OL898694.1 | 05-2021 | Brazil | | |
| OL898709.1 | 05-2021 | Brazil | | |
| OR167310.1 | 03-2023 | Brazil | | |
| KP164569.1 | 08-2014 | Brazil | 10.1186/s12916-015-0348-x | 25976325 |
| KP164568.1 | 05-2015 | Brazil | 10.1186/s12916-015-0348-x | 25976325 |
| KY055011.1 | 02-2016 | Brazil | | |
| MH823664.1 | 03-2017 | Brazil | 10.1007/s00705-019-04174-4 | 30729309 |
| MK518395.1 | 05-2017 | Brazil | 10.3390/v11121126 | 31817553 |
| OR167352.1 | 05-2023 | Brazil | | |
| OR167353.1 | 05-2023 | Brazil | | |
| OR167347.1 | 05-2023 | Brazil | | |
| OR167351.1 | 05-2023 | Brazil | | |
| OR167341.1 | 05-2023 | Brazil | | |
| OR167342.1 | 05-2023 | Brazil | | |
| OR167346.1 | 05-2023 | Brazil | | |
| OR167337.1 | 05-2023 | Brazil | | |
| OR167338.1 | 05-2023 | Brazil | | |

| | | |
|---|---|---|
| OR167339.1 | 05-2023 | Brazil |
| OR167336.1 | 04-2023 | Brazil |
| OR167335.1 | 04-2023 | Brazil |
| OR167334.1 | 04-2023 | Brazil |
| OR167333.1 | 04-2023 | Brazil |
| OR167350.1 | 04-2023 | Brazil |
| OR167332.1 | 04-2023 | Brazil |
| OR167348.1 | 04-2023 | Brazil |
| OR167354.1 | 04-2023 | Brazil |
| OR167340.1 | 04-2023 | Brazil |
| OR167331.1 | 04-2023 | Brazil |
| OR167330.1 | 04-2023 | Brazil |
| OR167308.1 | 04-2023 | Brazil |
| OR167349.1 | 04-2023 | Brazil |
| OR167327.1 | 03-2023 | Brazil |
| OR167328.1 | 03-2023 | Brazil |
| OR167307.1 | 03-2023 | Brazil |
| OR167343.1 | 03-2023 | Brazil |
| OR167322.1 | 03-2023 | Brazil |
| OR167306.1 | 03-2023 | Brazil |
| OR167318.1 | 03-2023 | Brazil |
| OQ865303.1 | 03-2023 | Brazil |
| OR167320.1 | 03-2023 | Brazil |
| OR167309.1 | 03-2023 | Brazil |
| OR167326.1 | 03-2023 | Brazil |
| OR167305.1 | 03-2023 | Brazil |
| OR167325.1 | 03-2023 | Brazil |
| OQ865308.1 | 03-2023 | Brazil |
| OQ865318.1 | 03-2023 | Brazil |
| OQ865313.1 | 03-2023 | Brazil |
| OR167321.1 | 03-2023 | Brazil |
| OR167329.1 | 03-2023 | Brazil |
| OR167302.1 | 03-2023 | Brazil |
| OR167317.1 | 03-2023 | Brazil |
| OR167303.1 | 03-2023 | Brazil |
| OR167304.1 | 03-2023 | Brazil |
| OR167313.1 | 03-2023 | Brazil |
| OR167323.1 | 03-2023 | Brazil |
| OQ865300.1 | 03-2023 | Brazil |
| OR063982.1 | 03-2023 | Brazil |
| OR063986.1 | 03-2023 | Brazil |
| OQ865315.1 | 03-2023 | Brazil |
| OR167300.1 | 03-2023 | Brazil |
| OR167301.1 | 03-2023 | Brazil |
| OQ865314.1 | 03-2023 | Brazil |

| | | |
|---|---|---|
| OQ865319.1 | 03-2023 | Brazil |
| OR064007.1 | 03-2023 | Brazil |
| OR063985.1 | 03-2023 | Brazil |
| OR167316.1 | 03-2023 | Brazil |
| OQ865324.1 | 03-2023 | Brazil |
| OQ865310.1 | 03-2023 | Brazil |
| OQ865316.1 | 02-2023 | Brazil |
| OR167319.1 | 02-2023 | Brazil |
| OR064004.1 | 02-2023 | Brazil |
| OR063983.1 | 02-2023 | Brazil |
| OR063979.1 | 02-2023 | Brazil |
| OR064003.1 | 02-2023 | Brazil |
| OR064006.1 | 02-2023 | Brazil |
| OR167311.1 | 02-2023 | Brazil |
| OR167312.1 | 02-2023 | Brazil |
| OR063980.1 | 02-2023 | Brazil |
| OR063981.1 | 02-2023 | Brazil |
| OR063987.1 | 02-2023 | Brazil |
| OR167315.1 | 02-2023 | Brazil |
| OR063984.1 | 02-2023 | Brazil |
| OR167314.1 | 02-2023 | Brazil |
| OR064015.1 | 02-2023 | Brazil |
| OR063988.1 | 02-2023 | Brazil |
| OR167299.1 | 02-2023 | Brazil |
| OR167324.1 | 02-2023 | Brazil |
| OR064001.1 | 02-2023 | Brazil |
| OR064002.1 | 02-2023 | Brazil |
| OR064000.1 | 02-2023 | Brazil |
| OR063978.1 | 02-2023 | Brazil |
| OR167296.1 | 02-2023 | Brazil |
| OR063996.1 | 02-2023 | Brazil |
| OR063999.1 | 02-2023 | Brazil |
| OR064005.1 | 02-2023 | Brazil |
| OR167297.1 | 02-2023 | Brazil |
| OR167298.1 | 02-2023 | Brazil |
| OR167295.1 | 02-2023 | Brazil |
| OR063994.1 | 02-2023 | Brazil |
| OQ865307.1 | 02-2023 | Brazil |
| OQ865323.1 | 02-2023 | Brazil |
| OR167294.1 | 02-2023 | Brazil |
| OR063998.1 | 02-2023 | Brazil |
| OR064014.1 | 02-2023 | Brazil |
| OR064017.1 | 02-2023 | Brazil |
| OR063997.1 | 02-2023 | Brazil |
| OR167293.1 | 02-2023 | Brazil |

| | | |
|---|---|---|
| OR064012.1 | 02-2023 | Brazil |
| OR064013.1 | 02-2023 | Brazil |
| OQ865309.1 | 02-2023 | Brazil |
| OR064029.1 | 02-2023 | Brazil |
| OR064030.1 | 02-2023 | Brazil |
| OQ865299.1 | 02-2023 | Brazil |
| OR064010.1 | 02-2023 | Brazil |
| OR063991.1 | 02-2023 | Brazil |
| OR063995.1 | 02-2023 | Brazil |
| OQ616991.1 | 02-2023 | Brazil |
| OQ616992.1 | 02-2023 | Brazil |
| OR063990.1 | 02-2023 | Brazil |
| OQ865311.1 | 02-2023 | Brazil |
| OQ865317.1 | 02-2023 | Brazil |
| OR063992.1 | 02-2023 | Brazil |
| OQ616990.1 | 02-2023 | Brazil |
| OQ865301.1 | 02-2023 | Brazil |
| OR063993.1 | 02-2023 | Brazil |
| OR064016.1 | 02-2023 | Brazil |
| OQ865321.1 | 01-2023 | Brazil |
| OQ865296.1 | 01-2023 | Brazil |
| OR064027.1 | 01-2023 | Brazil |
| OQ616989.1 | 01-2023 | Brazil |
| OQ865306.1 | 01-2023 | Brazil |
| OQ616988.1 | 01-2023 | Brazil |
| OQ616987.1 | 01-2023 | Brazil |
| OQ616986.1 | 01-2023 | Brazil |
| OQ616985.1 | 01-2023 | Brazil |
| OQ865322.1 | 01-2023 | Brazil |
| OQ865320.1 | 01-2023 | Brazil |
| OR064026.1 | 01-2023 | Brazil |
| OQ865298.1 | 01-2023 | Brazil |
| OR064011.1 | 01-2023 | Brazil |
| OR064025.1 | 01-2023 | Brazil |
| OR064024.1 | 01-2023 | Brazil |
| OR064008.1 | 01-2023 | Brazil |
| OR064009.1 | 01-2023 | Brazil |
| OR064023.1 | 01-2023 | Brazil |
| OR167291.1 | 01-2023 | Brazil |
| OR167292.1 | 01-2023 | Brazil |
| OR064022.1 | 01-2023 | Brazil |
| OR064020.1 | 01-2023 | Brazil |
| OR064019.1 | 01-2023 | Brazil |
| OR064021.1 | 01-2023 | Brazil |
| OR167289.1 | 01-2023 | Brazil |

| | | |
|---|---|---|
| OR167290.1 | 01-2023 | Brazil |
| OR064028.1 | 01-2023 | Brazil |
| OR064018.1 | 01-2023 | Brazil |
| OR063989.1 | 12-2022 | Brazil |
| OQ865312.1 | 12-2022 | Brazil |
| OQ865297.1 | 11-2022 | Brazil |
| OQ865305.1 | 11-2022 | Brazil |
| OQ865295.1 | 10-2022 | Brazil |
| OQ865304.1 | 10-2022 | Brazil |
| OQ759795.1 | 06-2022 | Brazil |
| OQ759799.1 | 06-2022 | Brazil |
| OQ760030.1 | 06-2022 | Brazil |
| OQ759686.1 | 06-2022 | Brazil |
| OQ759798.1 | 06-2022 | Brazil |
| OQ759972.1 | 06-2022 | Brazil |
| OQ759971.1 | 06-2022 | Brazil |
| OQ759973.1 | 06-2022 | Brazil |
| OR538602.1 | 06-2022 | Brazil |
| OQ759800.1 | 06-2022 | Brazil |
| OR538603.1 | 06-2022 | Brazil |
| OQ760029.1 | 06-2022 | Brazil |
| OQ759969.1 | 06-2022 | Brazil |
| OQ759970.1 | 06-2022 | Brazil |
| OQ759967.1 | 06-2022 | Brazil |
| OQ759968.1 | 06-2022 | Brazil |
| OQ759965.1 | 06-2022 | Brazil |
| OQ759815.1 | 06-2022 | Brazil |
| OQ759805.1 | 06-2022 | Brazil |
| OQ759804.1 | 06-2022 | Brazil |
| OQ759685.1 | 06-2022 | Brazil |
| OQ759966.1 | 06-2022 | Brazil |
| OQ759813.1 | 06-2022 | Brazil |
| OQ759811.1 | 06-2022 | Brazil |
| OQ759803.1 | 06-2022 | Brazil |
| OQ759810.1 | 06-2022 | Brazil |
| OQ759812.1 | 06-2022 | Brazil |
| OQ759814.1 | 06-2022 | Brazil |
| OQ759802.1 | 06-2022 | Brazil |
| OQ759963.1 | 06-2022 | Brazil |
| OQ759809.1 | 06-2022 | Brazil |
| OQ759808.1 | 06-2022 | Brazil |
| OQ759807.1 | 05-2022 | Brazil |
| OQ759684.1 | 05-2022 | Brazil |
| OQ759819.1 | 05-2022 | Brazil |
| OQ759806.1 | 05-2022 | Brazil |

| | | |
|---|---|---|
| OQ759824.1 | 05-2022 | Brazil |
| OQ759817.1 | 05-2022 | Brazil |
| OQ759820.1 | 05-2022 | Brazil |
| OQ759818.1 | 05-2022 | Brazil |
| OQ759826.1 | 05-2022 | Brazil |
| OQ759823.1 | 05-2022 | Brazil |
| OQ759816.1 | 05-2022 | Brazil |
| OQ760032.1 | 05-2022 | Brazil |
| OQ759822.1 | 05-2022 | Brazil |
| OQ759683.1 | 05-2022 | Brazil |
| OQ759827.1 | 05-2022 | Brazil |
| OQ759821.1 | 05-2022 | Brazil |
| OQ759801.1 | 05-2022 | Brazil |
| OQ759829.1 | 05-2022 | Brazil |
| OQ759830.1 | 05-2022 | Brazil |
| OQ759831.1 | 05-2022 | Brazil |
| OQ759832.1 | 05-2022 | Brazil |
| OQ759681.1 | 05-2022 | Brazil |
| OQ759828.1 | 05-2022 | Brazil |
| OQ759682.1 | 05-2022 | Brazil |
| OQ759825.1 | 05-2022 | Brazil |
| OQ759834.1 | 05-2022 | Brazil |
| OQ759833.1 | 05-2022 | Brazil |
| OQ759962.1 | 05-2022 | Brazil |
| OQ760026.1 | 05-2022 | Brazil |
| OQ760024.1 | 05-2022 | Brazil |
| OQ760031.1 | 05-2022 | Brazil |
| OQ760028.1 | 05-2022 | Brazil |
| OQ760021.1 | 05-2022 | Brazil |
| OQ760022.1 | 05-2022 | Brazil |
| OQ760023.1 | 05-2022 | Brazil |
| OQ759690.1 | 05-2022 | Brazil |
| OQ759688.1 | 05-2022 | Brazil |
| OQ759687.1 | 05-2022 | Brazil |
| OQ759964.1 | 05-2022 | Brazil |
| OQ760020.1 | 05-2022 | Brazil |
| OQ760017.1 | 05-2022 | Brazil |
| OQ760025.1 | 05-2022 | Brazil |
| OQ760014.1 | 05-2022 | Brazil |
| OQ760013.1 | 05-2022 | Brazil |
| OQ760027.1 | 05-2022 | Brazil |
| OQ760011.1 | 05-2022 | Brazil |
| OQ759961.1 | 05-2022 | Brazil |
| OQ759698.1 | 05-2022 | Brazil |
| OQ760015.1 | 05-2022 | Brazil |

| | | |
|---|---|---|
| OQ759960.1 | 05-2022 | Brazil |
| OQ760009.1 | 05-2022 | Brazil |
| OQ760019.1 | 05-2022 | Brazil |
| OQ760018.1 | 05-2022 | Brazil |
| OQ760010.1 | 05-2022 | Brazil |
| OQ760016.1 | 05-2022 | Brazil |
| OQ760008.1 | 05-2022 | Brazil |
| OQ759957.1 | 05-2022 | Brazil |
| OQ759956.1 | 05-2022 | Brazil |
| OQ760012.1 | 05-2022 | Brazil |
| OQ759689.1 | 04-2022 | Brazil |
| OQ759697.1 | 04-2022 | Brazil |
| OQ759696.1 | 04-2022 | Brazil |
| OQ759700.1 | 04-2022 | Brazil |
| OQ760007.1 | 04-2022 | Brazil |
| OQ759694.1 | 04-2022 | Brazil |
| OQ759958.1 | 04-2022 | Brazil |
| OQ759959.1 | 04-2022 | Brazil |
| OQ760006.1 | 04-2022 | Brazil |
| OQ759695.1 | 04-2022 | Brazil |
| OQ760005.1 | 04-2022 | Brazil |
| OQ760003.1 | 04-2022 | Brazil |
| OQ759988.1 | 04-2022 | Brazil |
| OQ760004.1 | 04-2022 | Brazil |
| OQ760001.1 | 04-2022 | Brazil |
| OQ759691.1 | 04-2022 | Brazil |
| OQ759692.1 | 04-2022 | Brazil |
| OQ759955.1 | 04-2022 | Brazil |
| OQ759954.1 | 04-2022 | Brazil |
| OQ759997.1 | 04-2022 | Brazil |
| OQ759953.1 | 04-2022 | Brazil |
| OQ759996.1 | 04-2022 | Brazil |
| OQ760002.1 | 04-2022 | Brazil |
| OQ759995.1 | 04-2022 | Brazil |
| OQ759999.1 | 04-2022 | Brazil |
| OQ759951.1 | 04-2022 | Brazil |
| OQ759998.1 | 04-2022 | Brazil |
| OQ759842.1 | 04-2022 | Brazil |
| OQ759948.1 | 04-2022 | Brazil |
| OQ759952.1 | 04-2022 | Brazil |
| OQ759699.1 | 04-2022 | Brazil |
| OQ759949.1 | 04-2022 | Brazil |
| OQ759947.1 | 04-2022 | Brazil |
| OQ759946.1 | 04-2022 | Brazil |
| OQ759941.1 | 04-2022 | Brazil |

| | | |
|---|---|---|
| OQ759945.1 | 04-2022 | Brazil |
| OQ759940.1 | 04-2022 | Brazil |
| OQ759943.1 | 04-2022 | Brazil |
| OQ759944.1 | 04-2022 | Brazil |
| OQ759992.1 | 04-2022 | Brazil |
| OQ759701.1 | 04-2022 | Brazil |
| OQ759938.1 | 04-2022 | Brazil |
| OQ759942.1 | 04-2022 | Brazil |
| OQ759939.1 | 04-2022 | Brazil |
| OQ759950.1 | 04-2022 | Brazil |
| OQ759937.1 | 04-2022 | Brazil |
| OQ759930.1 | 04-2022 | Brazil |
| OQ760000.1 | 04-2022 | Brazil |
| OQ759934.1 | 04-2022 | Brazil |
| OQ759936.1 | 04-2022 | Brazil |
| OQ759932.1 | 04-2022 | Brazil |
| OQ759931.1 | 04-2022 | Brazil |
| OQ759933.1 | 04-2022 | Brazil |
| OQ759935.1 | 04-2022 | Brazil |
| OQ759844.1 | 04-2022 | Brazil |
| OQ759927.1 | 04-2022 | Brazil |
| OQ759693.1 | 04-2022 | Brazil |
| OQ759924.1 | 04-2022 | Brazil |
| OQ759929.1 | 04-2022 | Brazil |
| OQ759923.1 | 04-2022 | Brazil |
| OQ759926.1 | 04-2022 | Brazil |
| OQ759922.1 | 04-2022 | Brazil |
| OQ759928.1 | 04-2022 | Brazil |
| OQ759915.1 | 04-2022 | Brazil |
| OQ759916.1 | 04-2022 | Brazil |
| OQ759925.1 | 04-2022 | Brazil |
| OQ616984.1 | 04-2022 | Brazil |
| OQ759914.1 | 04-2022 | Brazil |
| OQ759913.1 | 04-2022 | Brazil |
| OQ759918.1 | 04-2022 | Brazil |
| OQ759921.1 | 03-2022 | Brazil |
| OQ759917.1 | 03-2022 | Brazil |
| OQ759990.1 | 03-2022 | Brazil |
| OQ759991.1 | 03-2022 | Brazil |
| OQ759919.1 | 03-2022 | Brazil |
| OQ759989.1 | 03-2022 | Brazil |
| OQ759668.1 | 03-2022 | Brazil |
| OQ759920.1 | 03-2022 | Brazil |
| OQ759912.1 | 03-2022 | Brazil |
| OQ759911.1 | 03-2022 | Brazil |

| | | |
|---|---|---|
| OQ759896.1 | 03-2022 | Brazil |
| OQ759994.1 | 03-2022 | Brazil |
| OQ759895.1 | 03-2022 | Brazil |
| OQ759894.1 | 03-2022 | Brazil |
| OQ759893.1 | 03-2022 | Brazil |
| OQ759892.1 | 03-2022 | Brazil |
| OQ759664.1 | 03-2022 | Brazil |
| OQ759899.1 | 03-2022 | Brazil |
| OQ759677.1 | 03-2022 | Brazil |
| OQ759900.1 | 03-2022 | Brazil |
| OQ759666.1 | 03-2022 | Brazil |
| OQ759672.1 | 03-2022 | Brazil |
| OQ759665.1 | 03-2022 | Brazil |
| OQ759898.1 | 03-2022 | Brazil |
| OQ759660.1 | 03-2022 | Brazil |
| OQ759897.1 | 03-2022 | Brazil |
| OQ759910.1 | 03-2022 | Brazil |
| OQ759906.1 | 03-2022 | Brazil |
| OQ759661.1 | 03-2022 | Brazil |
| OQ759656.1 | 03-2022 | Brazil |
| OQ759909.1 | 03-2022 | Brazil |
| OQ759676.1 | 03-2022 | Brazil |
| OQ759908.1 | 03-2022 | Brazil |
| OQ759679.1 | 03-2022 | Brazil |
| OQ759903.1 | 03-2022 | Brazil |
| OQ759905.1 | 03-2022 | Brazil |
| OQ759902.1 | 03-2022 | Brazil |
| OQ759904.1 | 03-2022 | Brazil |
| OQ759901.1 | 03-2022 | Brazil |
| OQ759671.1 | 03-2022 | Brazil |
| OQ759891.1 | 03-2022 | Brazil |
| OQ759667.1 | 03-2022 | Brazil |
| OQ759889.1 | 03-2022 | Brazil |
| OQ759673.1 | 03-2022 | Brazil |
| OQ759840.1 | 03-2022 | Brazil |
| OQ759655.1 | 03-2022 | Brazil |
| OQ759680.1 | 03-2022 | Brazil |
| OQ759678.1 | 03-2022 | Brazil |
| OQ759888.1 | 03-2022 | Brazil |
| OQ616994.1 | 03-2022 | Brazil |
| OQ759993.1 | 03-2022 | Brazil |
| OQ759659.1 | 03-2022 | Brazil |
| OQ759663.1 | 03-2022 | Brazil |
| OQ759703.1 | 03-2022 | Brazil |
| OQ759657.1 | 03-2022 | Brazil |

| | | | | |
|---|---|---|---|---|
| OQ759670.1 | 03-2022 | Brazil | | |
| OQ616995.1 | 03-2022 | Brazil | | |
| OQ759843.1 | 03-2022 | Brazil | | |
| OQ759675.1 | 03-2022 | Brazil | | |
| OQ759839.1 | 03-2022 | Brazil | | |
| OQ759674.1 | 03-2022 | Brazil | | |
| OQ759669.1 | 03-2022 | Brazil | | |
| OQ759702.1 | 03-2022 | Brazil | | |
| OQ759841.1 | 03-2022 | Brazil | | |
| OP964986.1 | 03-2022 | Brazil | 10.1016/S2666-5247(23)00033-2 | 37031687 |
| OP964987.1 | 03-2022 | Brazil | 10.1016/S2666-5247(23)00033-2 | 37031687 |
| OP964988.1 | 03-2022 | Brazil | 10.1016/S2666-5247(23)00033-2 | 37031687 |
| OP964989.1 | 03-2022 | Brazil | 10.1016/S2666-5247(23)00033-2 | 37031687 |
| OP964990.1 | 03-2022 | Brazil | 10.1016/S2666-5247(23)00033-2 | 37031687 |
| OP964991.1 | 03-2022 | Brazil | 10.1016/S2666-5247(23)00033-2 | 37031687 |
| OP964992.1 | 03-2022 | Brazil | 10.1016/S2666-5247(23)00033-2 | 37031687 |
| OP964959.1 | 03-2022 | Brazil | 10.1016/S2666-5247(23)00033-2 | 37031687 |
| OQ759662.1 | 03-2022 | Brazil | | |
| OP964957.1 | 03-2022 | Brazil | 10.1016/S2666-5247(23)00033-2 | 37031687 |
| OP964958.1 | 03-2022 | Brazil | 10.1016/S2666-5247(23)00033-2 | 37031687 |
| OP964980.1 | 03-2022 | Brazil | 10.1016/S2666-5247(23)00033-2 | 37031687 |
| OP964981.1 | 03-2022 | Brazil | 10.1016/S2666-5247(23)00033-2 | 37031687 |
| OP964982.1 | 03-2022 | Brazil | 10.1016/S2666-5247(23)00033-2 | 37031687 |
| OP964983.1 | 03-2022 | Brazil | 10.1016/S2666-5247(23)00033-2 | 37031687 |
| OP964984.1 | 03-2022 | Brazil | 10.1016/S2666-5247(23)00033-2 | 37031687 |
| OP964985.1 | 03-2022 | Brazil | 10.1016/S2666-5247(23)00033-2 | 37031687 |
| OP964955.1 | 03-2022 | Brazil | 10.1016/S2666-5247(23)00033-2 | 37031687 |
| OP964977.1 | 03-2022 | Brazil | 10.1016/S2666-5247(23)00033-2 | 37031687 |
| OP964978.1 | 03-2022 | Brazil | 10.1016/S2666-5247(23)00033-2 | 37031687 |
| OP964979.1 | 03-2022 | Brazil | 10.1016/S2666-5247(23)00033-2 | 37031687 |
| OP964954.1 | 03-2022 | Brazil | 10.1016/S2666-5247(23)00033-2 | 37031687 |
| OP964956.1 | 03-2022 | Brazil | 10.1016/S2666-5247(23)00033-2 | 37031687 |
| OQ759837.1 | 03-2022 | Brazil | | |
| OP964960.1 | 03-2022 | Brazil | 10.1016/S2666-5247(23)00033-2 | 37031687 |
| OP964962.1 | 02-2022 | Brazil | 10.1016/S2666-5247(23)00033-2 | 37031687 |
| OP964976.1 | 02-2022 | Brazil | 10.1016/S2666-5247(23)00033-2 | 37031687 |
| OP964963.1 | 02-2022 | Brazil | 10.1016/S2666-5247(23)00033-2 | 37031687 |
| OP964961.1 | 02-2022 | Brazil | 10.1016/S2666-5247(23)00033-2 | 37031687 |
| OP964947.1 | 02-2022 | Brazil | 10.1016/S2666-5247(23)00033-2 | 37031687 |
| OP964948.1 | 02-2022 | Brazil | 10.1016/S2666-5247(23)00033-2 | 37031687 |
| OQ759886.1 | 02-2022 | Brazil | | |
| OP964950.1 | 02-2022 | Brazil | 10.1016/S2666-5247(23)00033-2 | 37031687 |
| OP964951.1 | 02-2022 | Brazil | 10.1016/S2666-5247(23)00033-2 | 37031687 |
| OP964952.1 | 02-2022 | Brazil | 10.1016/S2666-5247(23)00033-2 | 37031687 |
| OP964953.1 | 02-2022 | Brazil | 10.1016/S2666-5247(23)00033-2 | 37031687 |

| | | | | |
|---|---|---|---|---|
| OQ759838.1 | 02-2022 | Brazil | | |
| OP964949.1 | 02-2022 | Brazil | 10.1016/S2666-5247(23)00033-2 | 37031687 |
| OP964975.1 | 02-2022 | Brazil | 10.1016/S2666-5247(23)00033-2 | 37031687 |
| OP964967.1 | 02-2022 | Brazil | 10.1016/S2666-5247(23)00033-2 | 37031687 |
| OP964974.1 | 02-2022 | Brazil | 10.1016/S2666-5247(23)00033-2 | 37031687 |
| OP964972.1 | 02-2022 | Brazil | 10.1016/S2666-5247(23)00033-2 | 37031687 |
| OP964973.1 | 02-2022 | Brazil | 10.1016/S2666-5247(23)00033-2 | 37031687 |
| OQ759658.1 | 02-2022 | Brazil | | |
| OP964943.1 | 02-2022 | Brazil | 10.1016/S2666-5247(23)00033-2 | 37031687 |
| OP964944.1 | 02-2022 | Brazil | 10.1016/S2666-5247(23)00033-2 | 37031687 |
| OP964945.1 | 02-2022 | Brazil | 10.1016/S2666-5247(23)00033-2 | 37031687 |
| OP964946.1 | 02-2022 | Brazil | 10.1016/S2666-5247(23)00033-2 | 37031687 |
| OP964966.1 | 02-2022 | Brazil | 10.1016/S2666-5247(23)00033-2 | 37031687 |
| OP964968.1 | 02-2022 | Brazil | 10.1016/S2666-5247(23)00033-2 | 37031687 |
| OP964969.1 | 02-2022 | Brazil | 10.1016/S2666-5247(23)00033-2 | 37031687 |
| OP964970.1 | 02-2022 | Brazil | 10.1016/S2666-5247(23)00033-2 | 37031687 |
| OQ759884.1 | 02-2022 | Brazil | | |
| OQ759885.1 | 02-2022 | Brazil | | |
| OP964932.1 | 02-2022 | Brazil | 10.1016/S2666-5247(23)00033-2 | 37031687 |
| OP964933.1 | 02-2022 | Brazil | 10.1016/S2666-5247(23)00033-2 | 37031687 |
| OP964934.1 | 02-2022 | Brazil | 10.1016/S2666-5247(23)00033-2 | 37031687 |
| OP964935.1 | 02-2022 | Brazil | 10.1016/S2666-5247(23)00033-2 | 37031687 |
| OP964936.1 | 02-2022 | Brazil | 10.1016/S2666-5247(23)00033-2 | 37031687 |
| OP964937.1 | 02-2022 | Brazil | 10.1016/S2666-5247(23)00033-2 | 37031687 |
| OP964938.1 | 02-2022 | Brazil | 10.1016/S2666-5247(23)00033-2 | 37031687 |
| OP964939.1 | 02-2022 | Brazil | 10.1016/S2666-5247(23)00033-2 | 37031687 |
| OP964940.1 | 02-2022 | Brazil | 10.1016/S2666-5247(23)00033-2 | 37031687 |
| OP964941.1 | 02-2022 | Brazil | 10.1016/S2666-5247(23)00033-2 | 37031687 |
| OP964942.1 | 02-2022 | Brazil | 10.1016/S2666-5247(23)00033-2 | 37031687 |
| OQ616993.1 | 02-2022 | Brazil | | |
| OP964965.1 | 02-2022 | Brazil | 10.1016/S2666-5247(23)00033-2 | 37031687 |
| OQ759835.1 | 02-2022 | Brazil | | |
| OQ616996.1 | 02-2022 | Brazil | | |
| OQ759883.1 | 01-2022 | Brazil | | |
| OQ759781.1 | 01-2022 | Brazil | | |
| OQ759780.1 | 01-2022 | Brazil | | |
| OQ759782.1 | 01-2022 | Brazil | | |
| OQ759783.1 | 01-2022 | Brazil | | |
| OQ759796.1 | 01-2022 | Brazil | | |
| OQ759778.1 | 01-2022 | Brazil | | |
| OQ759784.1 | 01-2022 | Brazil | | |
| OQ759777.1 | 01-2022 | Brazil | | |
| OQ759758.1 | 01-2022 | Brazil | | |
| OQ759779.1 | 01-2022 | Brazil | | |
| OQ759776.1 | 01-2022 | Brazil | | |

| | | | | |
|---|---|---|---|---|
| KX496989.1 | 02-2016 | Colombia | | |
| KR559491.1 | 08-2014 | Colombia | | |
| MH329298.1 | 11-2014 | Colombia | | |
| MW656171.1 | 02-2015 | Colombia | | |
| MH329297.1 | 11-2014 | Colombia | | |
| MH329295.1 | 11-2014 | Colombia | | |
| MH329300.1 | 11-2014 | Colombia | | |
| MH359140.1 | 11-2014 | Colombia | | |
| MH359139.1 | 10-2014 | Colombia | | |
| MH329302.1 | 10-2014 | Colombia | | |
| LC259090.1 | 01-2015 | Colombia | 10.1093/jtm/tax072 | 29394382 |
| MH359142.1 | 10-2014 | Colombia | | |
| MH329293.1 | 11-2014 | Colombia | | |
| MH329299.1 | 11-2014 | Colombia | | |
| MH329294.1 | 10-2014 | Colombia | | |
| MH329303.1 | 10-2014 | Colombia | | |
| MH329296.1 | 09-2014 | Colombia | | |
| MN462646.1 | 05-2015 | Ecuador | | |
| MN462639.1 | 08-2015 | Ecuador | | |
| MN462655.1 | 07-2015 | Ecuador | | |
| MN462659.1 | 12-2015 | Ecuador | | |
| MN462644.1 | 05-2015 | Ecuador | | |
| MN462645.1 | 05-2015 | Ecuador | | |
| MN462662.1 | 08-2015 | Ecuador | | |
| MN462648.1 | 05-2015 | Ecuador | | |
| MN462641.1 | 05-2015 | Ecuador | | |
| MN462640.1 | 04-2015 | Ecuador | | |
| MN462642.1 | 11-2015 | Ecuador | | |
| MN462654.1 | 02-2015 | Ecuador | | |
| MN462658.1 | 04-2015 | Ecuador | | |
| MN462653.1 | 06-2015 | Ecuador | | |
| MN462647.1 | 05-2015 | Ecuador | | |
| MN462649.1 | 02-2015 | Ecuador | | |
| MN462651.1 | 06-2015 | Ecuador | | |
| MN462650.1 | 06-2015 | Ecuador | | |
| MN462660.1 | 08-2015 | Ecuador | | |
| KY435458.1 | 11-2014 | Guyana | 10.1093/ve/vex010 | 28480053 |
| KY435477.1 | 05-2014 | Guyana | 10.1093/ve/vex010 | 28480053 |
| KY435478.1 | 05-2014 | Guyana | 10.1093/ve/vex010 | 28480053 |
| KR559496.1 | 07-2014 | Guyana | | |
| KR559490.1 | 08-2014 | Guyana | | |
| MT038395.1 | 02-2016 | Paraguay | | |
| MT038393.1 | 01-2016 | Paraguay | | |
| MT038396.1 | 02-2016 | Paraguay | | |
| MT038394.1 | 02-2016 | Paraguay | | |

| | | |
|---|---|---|
| MT038397.1 | 01-2016 | Paraguay |
| MT038398.1 | 03-2016 | Paraguay |
| OQ775521.1 | 03-2023 | Paraguay |
| OQ775523.1 | 03-2023 | Paraguay |
| OQ775518.1 | 03-2023 | Paraguay |
| OQ775520.1 | 03-2023 | Paraguay |
| OQ775522.1 | 03-2023 | Paraguay |
| OQ775529.1 | 03-2023 | Paraguay |
| OQ775519.1 | 03-2023 | Paraguay |
| OQ775517.1 | 03-2023 | Paraguay |
| OQ775556.1 | 03-2023 | Paraguay |
| OQ775530.1 | 03-2023 | Paraguay |
| OQ775555.1 | 03-2023 | Paraguay |
| OQ775516.1 | 03-2023 | Paraguay |
| OQ775514.1 | 03-2023 | Paraguay |
| OQ775528.1 | 03-2023 | Paraguay |
| OQ775510.1 | 03-2023 | Paraguay |
| OQ775511.1 | 03-2023 | Paraguay |
| OQ775509.1 | 03-2023 | Paraguay |
| OQ775506.1 | 03-2023 | Paraguay |
| OQ775548.1 | 03-2023 | Paraguay |
| OQ775553.1 | 03-2023 | Paraguay |
| OQ775549.1 | 03-2023 | Paraguay |
| OQ775515.1 | 03-2023 | Paraguay |
| OQ775512.1 | 03-2023 | Paraguay |
| OQ775505.1 | 02-2023 | Paraguay |
| OQ775513.1 | 02-2023 | Paraguay |
| OQ775508.1 | 02-2023 | Paraguay |
| OQ775533.1 | 02-2023 | Paraguay |
| OQ775532.1 | 02-2023 | Paraguay |
| OQ775507.1 | 02-2023 | Paraguay |
| OQ775540.1 | 02-2023 | Paraguay |
| OQ775503.1 | 02-2023 | Paraguay |
| OQ775545.1 | 02-2023 | Paraguay |
| OQ775504.1 | 02-2023 | Paraguay |
| OQ775502.1 | 02-2023 | Paraguay |
| OQ775539.1 | 02-2023 | Paraguay |
| OQ775544.1 | 02-2023 | Paraguay |
| OQ775501.1 | 02-2023 | Paraguay |
| OQ775534.1 | 02-2023 | Paraguay |
| OQ775542.1 | 02-2023 | Paraguay |
| OQ775531.1 | 02-2023 | Paraguay |
| OQ775551.1 | 02-2023 | Paraguay |
| OQ775483.1 | 02-2023 | Paraguay |
| OQ775537.1 | 02-2023 | Paraguay |

| | | |
|---|---|---|
| OQ775538.1 | 02-2023 | Paraguay |
| OQ775552.1 | 02-2023 | Paraguay |
| OQ775554.1 | 02-2023 | Paraguay |
| OQ775567.1 | 02-2023 | Paraguay |
| OQ775471.1 | 02-2023 | Paraguay |
| OQ775468.1 | 02-2023 | Paraguay |
| OQ775469.1 | 02-2023 | Paraguay |
| OQ775499.1 | 02-2023 | Paraguay |
| OQ775467.1 | 02-2023 | Paraguay |
| OQ775479.1 | 02-2023 | Paraguay |
| OQ775497.1 | 02-2023 | Paraguay |
| OQ775465.1 | 02-2023 | Paraguay |
| OQ775566.1 | 02-2023 | Paraguay |
| OQ775464.1 | 02-2023 | Paraguay |
| OQ775478.1 | 02-2023 | Paraguay |
| OQ775462.1 | 02-2023 | Paraguay |
| OQ775565.1 | 02-2023 | Paraguay |
| OQ775463.1 | 02-2023 | Paraguay |
| OQ775460.1 | 02-2023 | Paraguay |
| OQ775564.1 | 02-2023 | Paraguay |
| OQ775459.1 | 02-2023 | Paraguay |
| OQ775498.1 | 02-2023 | Paraguay |
| OQ775496.1 | 02-2023 | Paraguay |
| OQ775493.1 | 02-2023 | Paraguay |
| OQ775495.1 | 02-2023 | Paraguay |
| OQ775457.1 | 02-2023 | Paraguay |
| OQ775455.1 | 02-2023 | Paraguay |
| OQ775458.1 | 02-2023 | Paraguay |
| OQ775456.1 | 02-2023 | Paraguay |
| OQ775453.1 | 01-2023 | Paraguay |
| OQ775452.1 | 01-2023 | Paraguay |
| OQ775473.1 | 01-2023 | Paraguay |
| OQ775449.1 | 01-2023 | Paraguay |
| OQ775448.1 | 01-2023 | Paraguay |
| OQ775492.1 | 01-2023 | Paraguay |
| OQ775494.1 | 01-2023 | Paraguay |
| OQ775461.1 | 01-2023 | Paraguay |
| OQ775474.1 | 01-2023 | Paraguay |
| OQ775561.1 | 01-2023 | Paraguay |
| OQ775563.1 | 01-2023 | Paraguay |
| OQ775562.1 | 01-2023 | Paraguay |
| OQ775446.1 | 01-2023 | Paraguay |
| OQ775451.1 | 01-2023 | Paraguay |
| OQ775443.1 | 01-2023 | Paraguay |
| OQ775490.1 | 01-2023 | Paraguay |

| | | |
|---|---|---|
| OQ775444.1 | 01-2023 | Paraguay |
| OQ775489.1 | 01-2023 | Paraguay |
| OQ775491.1 | 01-2023 | Paraguay |
| OQ775560.1 | 01-2023 | Paraguay |
| OQ775488.1 | 01-2023 | Paraguay |
| OQ775487.1 | 01-2023 | Paraguay |
| OQ775558.1 | 01-2023 | Paraguay |
| OQ775485.1 | 01-2023 | Paraguay |
| OQ775472.1 | 01-2023 | Paraguay |
| OQ775442.1 | 01-2023 | Paraguay |
| OQ775470.1 | 01-2023 | Paraguay |
| OQ775441.1 | 01-2023 | Paraguay |
| OQ775486.1 | 01-2023 | Paraguay |
| OQ775436.1 | 01-2023 | Paraguay |
| OQ775437.1 | 01-2023 | Paraguay |
| OQ775435.1 | 01-2023 | Paraguay |
| OQ775559.1 | 01-2023 | Paraguay |
| OQ775433.1 | 01-2023 | Paraguay |
| OQ567728.1 | 07-2022 | Paraguay |
| OQ567727.1 | 07-2022 | Paraguay |
| OQ775432.1 | 06-2022 | Paraguay |
| OQ775434.1 | 06-2022 | Paraguay |
| OQ775411.1 | 06-2022 | Paraguay |
| OQ775410.1 | 06-2022 | Paraguay |
| OQ775413.1 | 06-2022 | Paraguay |
| OQ775425.1 | 06-2022 | Paraguay |
| OQ775424.1 | 06-2022 | Paraguay |
| OQ775409.1 | 05-2022 | Paraguay |
| OQ775422.1 | 05-2022 | Paraguay |
| OQ775430.1 | 05-2022 | Paraguay |
| OQ775405.1 | 05-2022 | Paraguay |
| OQ775414.1 | 05-2022 | Paraguay |
| OQ775406.1 | 05-2022 | Paraguay |
| OQ775421.1 | 05-2022 | Paraguay |
| OQ775404.1 | 05-2022 | Paraguay |
| OQ775429.1 | 05-2022 | Paraguay |
| OQ775407.1 | 05-2022 | Paraguay |
| OQ775403.1 | 05-2022 | Paraguay |
| OQ775427.1 | 05-2022 | Paraguay |
| OQ775408.1 | 05-2022 | Paraguay |
| OQ775420.1 | 05-2022 | Paraguay |
| OQ775423.1 | 05-2022 | Paraguay |
| OQ775401.1 | 05-2022 | Paraguay |
| OQ775415.1 | 05-2022 | Paraguay |
| OQ775402.1 | 05-2022 | Paraguay |

| | | | | |
|---|---|---|---|---|
| OQ775399.1 | 05-2022 | Paraguay | | |
| OQ775419.1 | 05-2022 | Paraguay | | |
| OQ775397.1 | 04-2022 | Paraguay | | |
| OQ775428.1 | 04-2022 | Paraguay | | |
| OQ775400.1 | 04-2022 | Paraguay | | |
| OQ775396.1 | 04-2022 | Paraguay | | |
| OQ567723.1 | 04-2022 | Paraguay | 10.3201/eid2909.230523 | 37488810 |
| OQ775412.1 | 04-2022 | Paraguay | | |
| OQ775418.1 | 04-2022 | Paraguay | | |
| OQ567722.1 | 04-2022 | Paraguay | 10.3201/eid2909.230523 | 37488810 |
| OQ775417.1 | 04-2022 | Paraguay | | |
| OQ567724.1 | 04-2022 | Paraguay | 10.3201/eid2909.230523 | 37488810 |
| OQ775416.1 | 04-2022 | Paraguay | | |
| OQ567726.1 | 04-2022 | Paraguay | | |
| OQ567725.1 | 04-2022 | Paraguay | 10.3201/eid2909.230523 | 37488810 |
| OQ775398.1 | 04-2022 | Paraguay | | |
| OQ775426.1 | 04-2022 | Paraguay | | |
| OQ775394.1 | 04-2022 | Paraguay | | |
| OQ775395.1 | 04-2022 | Paraguay | | |
| KY435463.1 | 08-2014 | Suriname | 10.1093/ve/vex010 | 28480053 |
| KY435456.1 | 08-2014 | Suriname | 10.1093/ve/vex010 | 28480053 |
| OR360566.1 | 03-2023 | Uruguay | | |
| OR360567.1 | 02-2023 | Uruguay | | |
| OR360568.1 | 03-2023 | Uruguay | | |
| OR360569.1 | 02-2023 | Uruguay | | |
| OR360570.1 | 03-2023 | Uruguay | | |
| OR360571.1 | 04-2023 | Uruguay | | |
| OR360573.1 | 04-2023 | Uruguay | | |
| OR360574.1 | 04-2023 | Uruguay | | |
| OR360575.1 | 04-2023 | Uruguay | | |
| OR360577.1 | 04-2023 | Uruguay | | |
| OR360578.1 | 04-2023 | Uruguay | | |
| OR360579.1 | 04-2023 | Uruguay | | |
| OR360580.1 | 04-2023 | Uruguay | | |
| OR360581.1 | 04-2023 | Uruguay | | |
| OR360582.1 | 04-2023 | Uruguay | | |
| OR360583.1 | 04-2023 | Uruguay | | |
| OR360584.1 | 04-2023 | Uruguay | | |
| OR360585.1 | 05-2023 | Uruguay | | |
| OR360586.1 | 04-2023 | Uruguay | | |
| OR360587.1 | 05-2023 | Uruguay | | |
| OR360588.1 | 05-2023 | Uruguay | | |
| OR360589.1 | 05-2023 | Uruguay | | |
| OR360590.1 | 05-2023 | Uruguay | | |
| OR360591.1 | 05-2023 | Uruguay | | |

| | | |
|---|---|---|
| OR360592.1 | 05-2023 | Uruguay |
| OR360593.1 | 05-2023 | Uruguay |
| OR360594.1 | 05-2023 | Uruguay |
| OR360595.1 | 05-2023 | Uruguay |
